# Dissecting the pleiotropic genetic architecture linking telomere biology to chronic respiratory diseases and lung function

**DOI:** 10.1101/2025.07.09.25331189

**Authors:** Pengwei Zhang, Leilei Zheng, Jun Qiao, Yuzhe Fan, Siyuan Chang, Yuxin Ning, Yixuan Yang, Mengting Liu, Xinying Hao, Jinyu Gao, Yuliang Feng

## Abstract

**Background:** Leukocyte telomere length (LTL) has been implicated in aging and age-related diseases, including chronic respiratory diseases (CRDs). However, the extent and mechanisms of shared genetic architecture between LTL and respiratory health indicators (such as CRDs and lung function parameters) remain incompletely understood.

**Methods:** We first systematically characterized the genetic correlations and genetic overlaps between LTL and multiple respiratory health indicators. We then performed horizontal pleiotropy analysis by integrating SNP-level functional annotation, gene mapping, and pathway analysis to identify candidate pleiotropic loci, genes, and shared biological pathways. Finally, we assessed the causal relationships among these trait pairs using the latent causal variable (LCV) and MRlap.

**Results:** There was extensive genetic overlap between LTL and respiratory health indicators, regardless of whether these trait pairs had significant genetic associations. We identified 27,885 candidate pleiotropic loci and 82 pleiotropic genes. Notably, five key genes, such as *STN1*, *MPHOSPH9*, *MTRFR*, *MAX*, and *UCKL1*, showed significant pleiotropic effects in multiple phenotype pairs and mediated different patterns of genetic associations. Pathway enrichment analysis of these pleiotropic genes highlighted the close association of specific trait pairs with RNA metabolism and telomere maintenance. Finally, vertical pleiotropy analysis revealed negative causal associations of LTL-idiopathic pulmonary fibrosis and chronic obstructive pulmonary disease-LTL.

**Conclusions:** Our findings reinforce the roles of telomere biology and RNA processing in the pathogenesis of chronic lung diseases and support a shared genetic basis for common molecular pathways.

**Graphical Abstract: Analysis flow chart of LTL and seven respiratory health indicators:** 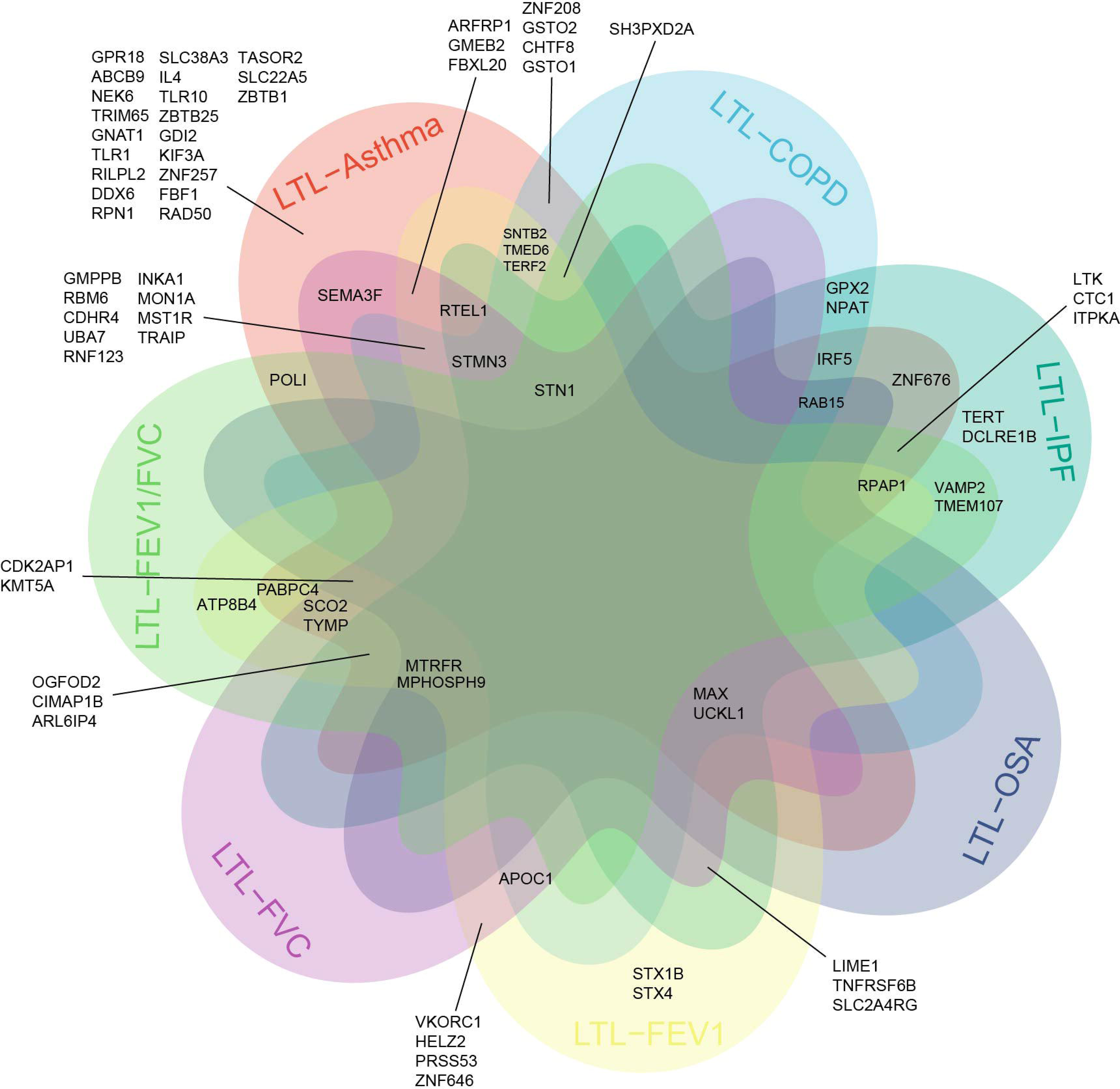

We used large-scale genome-wide association study (GWAS) summary statistics from individuals of European ancestry to conduct a comprehensive pleiotropy analysis of leukocyte telomere length (LTL) and seven respiratory health indicators. First, we systematically assessed genome-wide genetic correlation, global and local genetic overlap, and local genetic correlation across these phenotype pairs, revealing a shared genetic architecture. Recognizing that such associations may arise from vertical or horizontal pleiotropy, we next focused on capturing horizontal pleiotropy. Using multiple complementary statistical genetics methods, we integrated multidimensional evidence—spanning shared single-nucleotide polymorphisms (SNPs), genes, and molecular pathways—to characterize the genetic interconnections between LTL and respiratory traits. Finally, we performed causal inference analyses to evaluate the role of vertical pleiotropy, estimating the directional effects of LTL on respiratory health indicators. Together, this multi-layered pleiotropy analysis delineates a shared genetic landscape between LTL and respiratory health, offering novel insights into the mechanisms of lung aging and disease progression. LTL, leukocyte telomere length; COPD, chronic obstructive pulmonary disease; IPF, idiopathic pulmonary fibrosis; OSA, sleep apnoea; FEV1, forced expiratory volume in 1 second; FVC, forced vital capacity.

**take home:**

- The first study to comprehensively explore the common genetic architecture between LTL and respiratory health indicators (CRDs and LF parameters)
- 12q24.31 (*MPHOSPH9*, *MTRFR*), 14q23.3 (*MAX*), 20q13.33 (*UCKL1*) and 10q24.33 (*STN1*) are key pleiotropic loci and genes between LTL and respiratory health indicators

Genetically predicted LTL is causally associated with increased risk of IPF

## Introduction

Chronic respiratory diseases (CRDs) have become a growing public health concern, with increasing rates of morbidity and mortality worldwide[1][2][3]. Among them, chronic obstructive pulmonary disease (COPD) is particularly severe, ranking as the third leading cause of death globally and accounting for approximately 3.3 million deaths annually[4]. Smoking and indoor air pollution are well-established risk factors for various CRDs, and recent research further underscores aging as a critical driver of disease progression[5][6]. Lung function (LF) naturally declines with age due to peripheral airway narrowing, reduced lung elasticity, and enlarged alveolar spaces[7]. At the cellular level, aging is characterized by cellular senescence, which is an irreversible arrest of the cell cycle. This process impairs tissue repair and remodeling and alters the secretion of pro-inflammatory factors, contributing to low-grade chronic inflammation[8]. The accumulation of senescent cells in the lung may lead to small airway fibrosis, alveolar cell loss, and progressive pulmonary fibrosis. Notably, while aging may drive COPD progression, recent studies suggest a bidirectional relationship. Telomere shortening in circulating leukocytes of COPD patients indicates that COPD itself may accelerate biological aging[9][10]. Given the critical role of leukocyte telomere length (LTL) as a “molecular clock” of biological aging, its progressive attrition may be key to understanding the pathophysiology of CRDs and the associated decline in LF. Although clinical and epidemiological studies have provided preliminary evidence supporting this association, the precise mechanistic role of LTL in CRDs and LF decline remains to be fully elucidated.

Recent research has shown that individuals with inherited mutations in telomerase and other LTL-regulating genes exhibit up to a 90% increased risk of developing certain CRDs, such as idiopathic pulmonary fibrosis (IPF)[11]. Large-scale genome-wide association studies (GWAS) have identified common genetic variants that influence both LTL and susceptibility to CRDs[12][13]. For example, mutations in the *RTEL1* gene, a key regulator of LTL, have been linked to increased IPF risk, underscoring a genetic connection between LTL and respiratory disease susceptibility[13]. This genetic overlap may be attributed to pleiotropy, where a single genetic variant affects multiple traits. Pleiotropy can be categorized as vertical or horizontal: vertical pleiotropy occurs when a variant influences one trait through its effect on another, while horizontal pleiotropy refers to a variant affecting two traits independently via separate biological pathways. Mendelian randomization (MR) is a powerful tool for detecting vertical pleiotropy and inferring causal relationships. For example, Wang et al.[14] used MR to demonstrate that longer LTL may reduce the risk of COPD by modulating inflammatory markers such as C-reactive protein, white blood cell count, and neutrophil count. However, contrasting findings have emerged; another MR study reported that genetically shorter LTL increases IPF risk but has no significant effect on COPD[15]. These discrepancies may stem from differences in disease pathophysiology, genetic background, or the inherent limitations of MR in fully capturing complex pleiotropic effects. Recognizing the complexity of these genetic interactions, recent studies have turned to using horizontal pleiotropy to uncover underlying genetic mechanisms. For example, Gong et al.[16] demonstrated a shared genetic basis for gastrointestinal and psychiatric disorders by sequentially identifying common SNPs, genes, and biological pathways. Therefore, a systematic investigation of pleiotropic relationships between LTL and respiratory health indicators (such as CRDs and LF parameters), is essential to unravel their shared genetic architecture and deepen our understanding of respiratory aging and disease mechanisms.

In this study, we leveraged large-scale data from the GWAS consortium to explore the genetic correlations and overlaps between LTL and respiratory health indicators (Asthma, COPD, IPF, obstructive sleep apnea [OSA], forced vital capacity [FVC], forced expiratory volume in 1 second [FEV1], and the FEV1/FVC ratio). We first identified genetic variants contributing to the shared genetic architecture of LTL and each respiratory health indicator at the SNP level and performed pairwise co-localization analyses. Subsequently, we conducted functional annotation and genetic mapping of co-localized loci to identify candidate pleiotropic genes. Furthermore, we elucidated key biological pathways potentially linking LTL with CRDs and LF parameters. Finally, we applied MR analysis to infer potential causal relationships between LTL and respiratory health indicators. Overall, our study provides novel insights into how accelerated aging, as indicated by shortened LTL, may influence CRD pathogenesis and individual differences in LF.

## Methods

### GWAS data ethics and quality control

Given the current lack of well-powered GWAS in non-European populations, we restricted our analyses to individuals of European ancestry. We sourced GWAS summary statistics for LTL and respiratory health indicators from publicly available datasets. Specifically, GWAS summary statistics for LTL were obtained from a large-scale meta-analysis of the UK Biobank, comprising 472,174 participants of European ancestry[17]. For CRDs, data for Asthma and COPD were obtained from the Global Biobank Meta-analysis Initiative, which included 121,940 Asthma patients and 1,254,131 controls[18], and 58,559 COPD patients and 937,358 controls, respectively. GWAS summary statistics for IPF were sourced from the largest available meta-analysis, which combined five studies and included 4,125 cases and 20,464 controls of European ancestry[19]. For OSA, we used GWAS summary data from the FinnGen study, consisting of 33,423 OSA patients and 307,648 controls of European ancestry. Finally, GWAS summary statistics for three LF parameters—FVC, FEV1, and the FEV1/FVC ratio—were derived from the UK Biobank dataset, including 321,047 European-ancestry participants[20]. All GWAS studies utilized in this analysis had been approved by the respective institutional ethics committees. Detailed information on the datasets used is provided in Supplementary Table 1.

### Statistical Analysis

Graphical Abstract presents the workflow for this study. All analyses excluded SNPs in the major histocompatibility complex region (chr6: 25–35 Mb) and restricted analyses to biallelic SNPs with minor allele frequency larger than 0.01. Details of the statistical analyses are provided in the eMethods in the Supplementary Material.

## Result

### Estimation of global and local genetic correlations

We estimated the genome-wide genetic correlations (*r_g_*) between LTL and respiratory health indicators using bivariate linkage disequilibrium score regression (LDSC). Five trait pairs showed statistically significant genome-wide *r_g_* after Bonferroni correction (*P* < 7.14 × 10^−3^). Specifically, LTL exhibited negative *r_g_* with Asthma (*r_g_* = −0.122, SE = 0.022), COPD (*r_g_* = −0.221, SE =0.031), and IPF (*r_g_* = −0.315, SE = 0.058), and positive *r_g_* with FEV1 (*r_g_* = 0.065, SE = 0.020) and FVC (*r_g_* = 0.056, SE =0.020) (Supplementary Table 2).

Unlike LDSC, which captures genome-wide averaged pleiotropic effects, Local Analysis of [Co]Variant Annotation (LAVA) estimates local-*r_g_s* within specific genomic regions. Using LAVA, we identified 661 significant regions associated with LTL and 4,628 with respiratory health indicators (*P* < 1 × 10^−4^) (Supplementary Table 3). We then tested local-*r_g_s* across 1,822 genomic partitions and found 430 regions with nominally significant correlations (*P* < 0.05) (Figure 1A and Supplementary Table 4). A total of 196 genomic regions remained significant across all trait pairs after FDR correction (FDR < 0.05), highlighting the robustness of the local correlation patterns. Notably, although the global *r_g_* between LTL and OSA was not significant, we identified 11 genomic regions showing significant local-*r_g_s* (5 positive and 6 negative). This suggests opposing effects at different loci that may cancel out in genome-wide analyses. Importantly, the direction of the local-*r_g_s* in the five significant trait pairs (LTL – Asthma, LTL – COPD, LTL – IPF, LTL – FEV1, and LTL – FVC) was concordant with the genome-wide *r_g_* estimates from LDSC. In conclusion, local-*r_g_s* provide a finer-scale view of the genetic relationships between LTL and respiratory health indicators, revealing complex etiological patterns not captured by genome-wide analyses.

**Figure 1:**
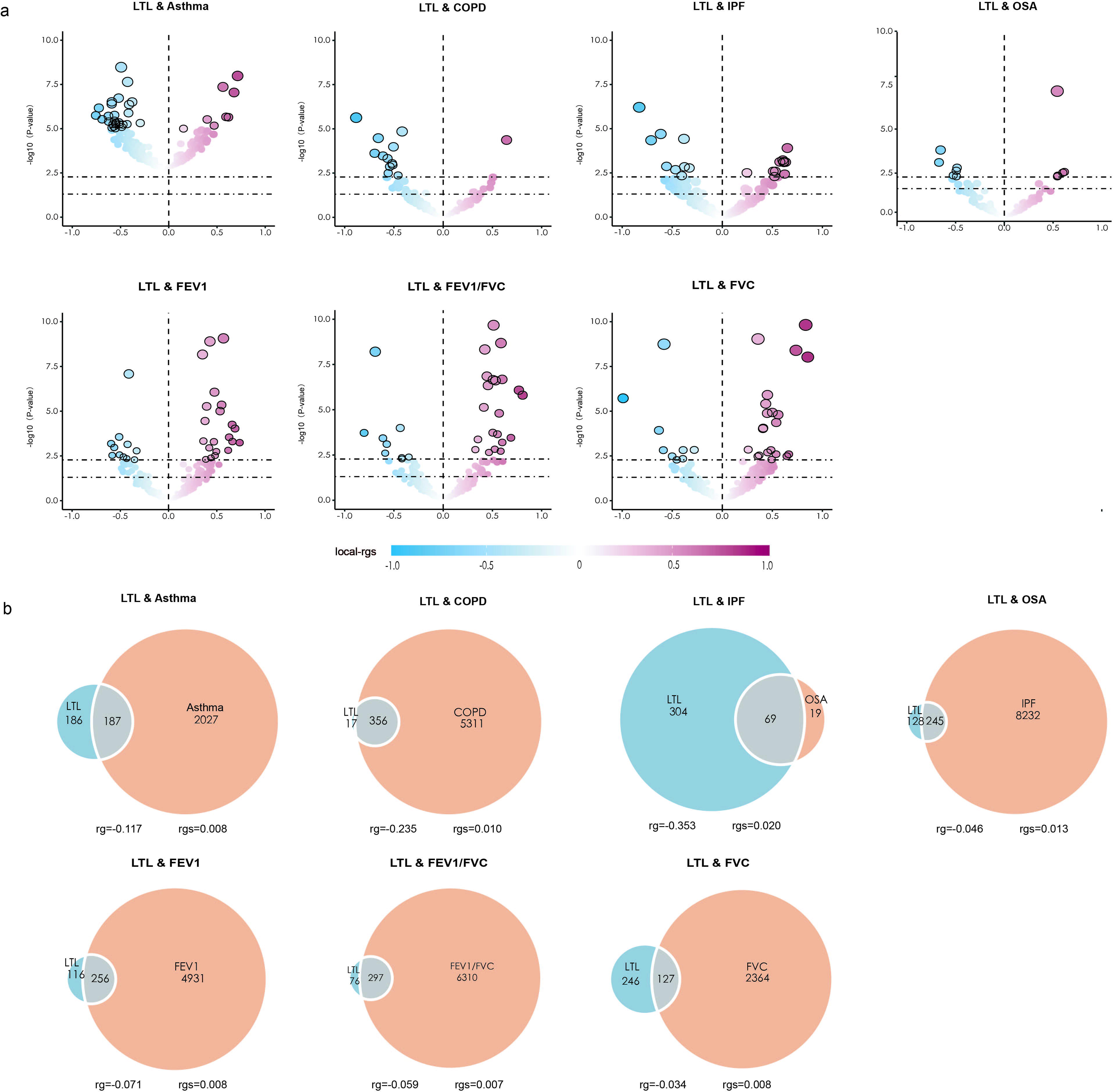
Local genetic correlations and genetic overlap between leukocyte telomere length and seven key respiratory health indicators. A) LAVA volcano plots showing local genetic correlation coefficients (local-rgs, y-axis) for LTL and major respiratory health indicatorsversus with −log10 p-values for pairwise analysis of each locus. Larger points with black circles indicate loci that are significantly associated after FDR correction (FDR < 0.05). Blue indicates negative correlations, red indicates positive correlations. LAVA-estimated local-rgs are shown on a blue-red scale. B) Venn diagram showing shared and disease-specific “causal” variants between LTL (blue) and respiratory health indicators (orange) estimated by MiXeR. Numbers indicate the estimated number of variants per phenotype that explain 90% of the SNP heritability for each phenotype. The size of the circle indicates the degree of polygenicity for each phenotype, with larger circles indicating stronger polygenicity. Polygenic overlap is indicated in gray.

### Quantification of genetic overlap using mixture modeling approach (MiXeR)

Although no globally consistent genetic associations were observed between LTL and specific respiratory health indicators across the genome, pleiotropic signals may still exist at specific loci. To further explore this possibility, we applied MiXeR to estimate the number of “causal” variants for each trait (univariate analysis) and to quantify the shared and unique variants between trait pairs (bivariate analysis).

We found that LTL is less polygenic, with only 373 causal variants explaining 90% of its SNP-based heritability (h2SNP) (SD = 0.026). Among respiratory health indicators, IPF showed the lowest polygenicity (N = 87, SD = 22), followed by Asthma (N = 2,214, SD = 131) and FEV1/FVC (N = 2,491, SD = 64). In contrast, COPD and FEV1 had higher polygenicity (5,188 – 5,667 variants), while FVC (N = 6,607, SD = 116) and OSA (N = 8,476, SD = 820) exhibited the highest levels of polygenicity (Supplementary Table 5A). Bivariate MiXeR analysis revealed distinct patterns of genetic overlap between LTL and various respiratory health indicators (Figure 1B, Supplementary Figure 1, and Supplementary Table 5B). The overlap between LTL and COPD was particularly striking. A total of 356 shared variants were identified, representing 95.56% of LTL’s causal variants but only 6.28% of those for COPD. This asymmetry reflects LTL’s lower polygenicity and the greater complexity of COPD’ s genetic architecture. The Dice coefficient of 0.118 (SD = 0.008) further supported this overlap, aligning with the relatively strong negative genome-wide genetic correlation observed via LDSC. In contrast, although 65.68% of LTL’s causal variants were also implicated in OSA, the genome-wide *r_g_* between LTL and OSA was close to zero. This paradoxical finding suggests that while many variants are shared, the directions of their effects are largely inconsistent. Indeed, only about 36% of the shared variants had concordant effects on both traits. Moreover, OSA exhibited substantially higher polygenicity than LTL, with an estimated 8,476 causal variants compared to 373 for LTL. Of the shared genetic architecture, only 245 variants were common to both traits, while the majority remained trait-specific (8,232 unique to OSA and 128 unique to LTL).

Similar patterns of partial overlap and mixed-effect directions were observed between LTL and other respiratory health indicators, except IPF, where shared polygenicity was minimal. Collectively, these findings highlight the complex polygenic architecture underlying LTL and its associations with respiratory health indicators, suggesting shared molecular mechanisms despite heterogeneous genetic effects.

### Identification and functional annotation of shared loci

Given the extensive genetic overlap between LTL and respiratory health indicators, we applied pleiotropy analysis under a composite null hypothesis (PLACO) to identify shared risk SNPs that may mediate horizontal pleiotropy across these traits. This analysis identified a total of 27,885 SNPs with evidence of pleiotropic effects across LTL and respiratory health indicators. To gain insight into the potential functional relevance of these variants, we conducted functional annotation using Functional Mapping and Annotation (FUMA), which revealed 329 independent genomic risk loci spanning 131 unique chromosomal regions (Figure 2, Supplementary Figure 2, and Supplementary Table 6). Notably, 83 of these regions were shared across multiple trait pairs, and several (such as 12q24.31, 20q13.33, and 3q26.2) emerged as recurrent pleiotropic loci across all examined trait combinations. Furthermore, loci at 14q23.3 and 10q24.33 were found to influence up to six different trait pairs, further underscoring their broad pleiotropic potential. Previous studies have implicated the 12q24.31 region in airway responsiveness, supporting its possible involvement in both CRDs and LF parameters[21].

**Figure 2:**
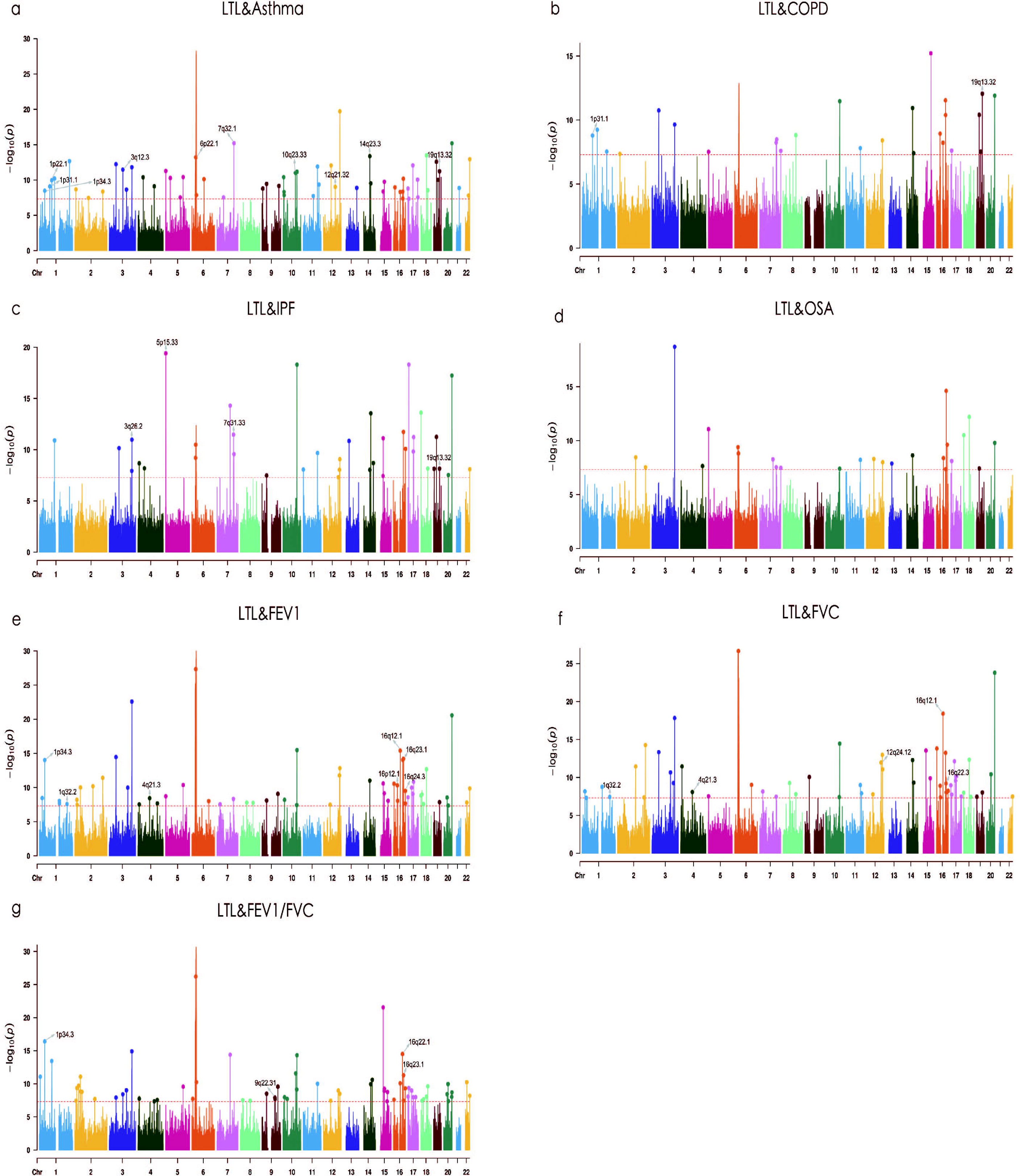
Manhattan plots of leukocyte telomere length and sseven key respiratory health indicators. The x-axis reflects chromosomal position and the y-axis reflects the negative log10-transformed P-value for each SNP. The horizontal dashed red line indicates a genome-wide significant *P*-value of −log_10_ (5×10^−8^). Independent genome-wide significant associations with the smallest *P*-values (Top lead SNP) are circled with colored circles. Only SNPs shared across all summary statistics are included. Labels are chromosomal regions where genomic risk loci with strong colocalization evidence (PP.H4 > 0.7) are located.

Functional classification using ANNOVAR indicated that among the SNPs located within the pleiotropic loci, 176 (53.50%) were intronic, 102 (31.00%) were intergenic, and only 14 (4.26%) were exonic. The predominance of intronic and intergenic variants suggests that regulatory elements may play a critical role in mediating shared genetic influences between LTL and respiratory health indicators.. For instance, we identified a locus at the *MPHOSPH9* gene (lead SNP rs10744151), located within an intronic region on 12q24.31, which was jointly associated with LTL and Asthma. This SNP also showed significant eQTL associations in multiple tissues, including lung (*P* = 3.04 × 10^−6^), sun-exposed skin (lower leg; *P* = 9.83 × 10^−11^), non – sun-exposed skin (suprapubic, *P* = 6.18×10^−4^), and whole blood (*P* = 2.28×10^−4^) (Supplementary Table 8). Despite these strong regulatory signals, no prior studies have reported a link between *MPHOSPH9* and LTL-Asthma, suggesting a potentially novel pleiotropic role for this gene.

Interestingly, the allelic directions of association at the top SNPs from these 329 pleiotropic loci were heterogeneous (Supplementary Table 6). Specifically, 68 SNPs (20.67%) conferred increased risk for both traits in a given pair, 91 SNPs (27.66%) conferred decreased risk for both, while 170 SNPs (51.67%) exhibited opposite directions of association. These mixed patterns highlight the complex nature of pleiotropic relationships between LTL and respiratory phenotypes. Collectively, these findings suggest that pleiotropic variants, particularly those in non-coding regulatory regions, may contribute to shared genetic susceptibility across LTL and respiratory health indicators, warranting further functional validation.

Among these pleiotropic variants, we identified 31 SNPs, corresponding to 24 unique loci, with potentially deleterious effects as indicated by high Combined Annotation-Dependent Depletion (CADD) scores (>12.37) (Supplementary Table 6). Notably, rs2277339, a non-synonymous variant in the *PRIM1* gene, exhibited the highest CADD score of 28.2, suggesting a substantial potential for functional disruption. Furthermore, 16 SNPs (13 unique) were predicted to affect transcription factor binding, with the strongest functional evidence supported by a RegulomeDB score of 1f. These included variants such as rs2271960 and rs4902352, which may play critical regulatory roles and thereby contribute to the pleiotropic associations between LTL and respiratory health indicators.

### Colocalization analysis

To further elucidate potential causal variants underlying these pleiotropic loci, we conducted colocalization analysis for each locus across trait pairs (Supplementary Figure 3 and Supplementary Table 6). Our results revealed that 93 loci (28.27%) had a posterior probability for hypothesis 3 (PPH3) greater than 0.7, indicating that different causal variants are likely driving the associations at these loci. A total of 32 loci (9.73%) exhibited a posterior probability for hypothesis 4 (PPH4) greater than 0.7, suggesting that a shared causal variant may underlie the associations for both traits in each pair. Among these, we identified 20 top SNPs as putative shared causal variants for the corresponding trait pairs. Of particular interest, rs429358, a well-characterized variant located in the *APOE* gene at 19q13.32, was consistently colocalized across three trait pairs: TL-Asthma, TL-COPD, and TL-IPF. This SNP, which is part of the *APOE* ε2/ε3/ε4 haplotype known to influence lipid metabolism and neurodegeneration[22][23][24], may also play a central role in mediating telomere biology and chronic respiratory outcomes. Another notable colocalized variant was rs762810, located in the 14q23.3 region and mapped to the *MAX* gene, which was specifically associated with both LTL and Asthma. These findings suggest that such variants may be key genetic contributors to the observed pleiotropy between LTL and respiratory health indicators, potentially through regulatory and gene expression mechanisms.

### Prioritization of shared putatively functional genes

To further elucidate the functional implications of the pleiotropic loci jointly associated with LTL and respiratory health indicators, we employed two complementary gene-mapping strategies to identify candidate target genes. The first approach utilized positional gene mapping based on proximity using multi-marker analysis of genomic annotation (MAGMA) analysis, while the second integrated tissue-specific cis-eQTL data through eQTL-informed MAGMA (e-MAGMA), enabling the assignment of distal regulatory variants to their potential target genes.

MAGMA analysis identified 382 significant gene-level associations across pleiotropic loci, corresponding to 122 unique genes (Figure 3 and Supplementary Table 10). Of these, 85 genes (69.67%) were associated with more than one trait pair, indicating widespread genetic pleiotropy. For instance, *SH3PXD2A*, located at 10q24.33, was significantly associated with seven different trait pairs. Fourteen genes were shared across six trait pairs, including clusters within key chromosomal regions such as 20q13.33 (7 genes), 12q24.31 (4 genes), and 3q26.2 (e.g., *MECOM*). Notably, *SH3PXD2A* (also known as *TKS5)* encodes a cytoskeletal regulator involved in cell migration and epithelial remodeling[[25]] and has been implicated as a susceptibility locus in multiple CRDs[26][27][28]. *MECOM,* located at 3q26.2, has been identified in several GWAS as a risk locus for COPD, and LF, and regulates inflammatory transcriptional programs[29][30][31]. Although their direct role in telomere biology remains unclear, these genes may represent molecular links bridging LTL regulation and respiratory disease. Several genes mapped by MAGMA are also known to be involved in telomere maintenance. For example, *STN1* (located at 10q24.33) plays a critical role in protecting telomeric and subtelomeric regions during DNA replication[32][33]. Overexpression of *STN1* can restore telomere length and enhance cell viability under stress, and its mutations have been associated with pulmonary fibrosis, emphysema, and pulmonary vascular disease[34]. Among the identified genes, 39 were newly associated with LTL, and 225 were newly linked to respiratory health indicators (Supplementary Table 10). Importantly, 95.81% of MAGMA-mapped genes were independently validated through FUMA positional mapping, providing strong support for their assignment (Supplementary Table 8).

**Figure 3:**
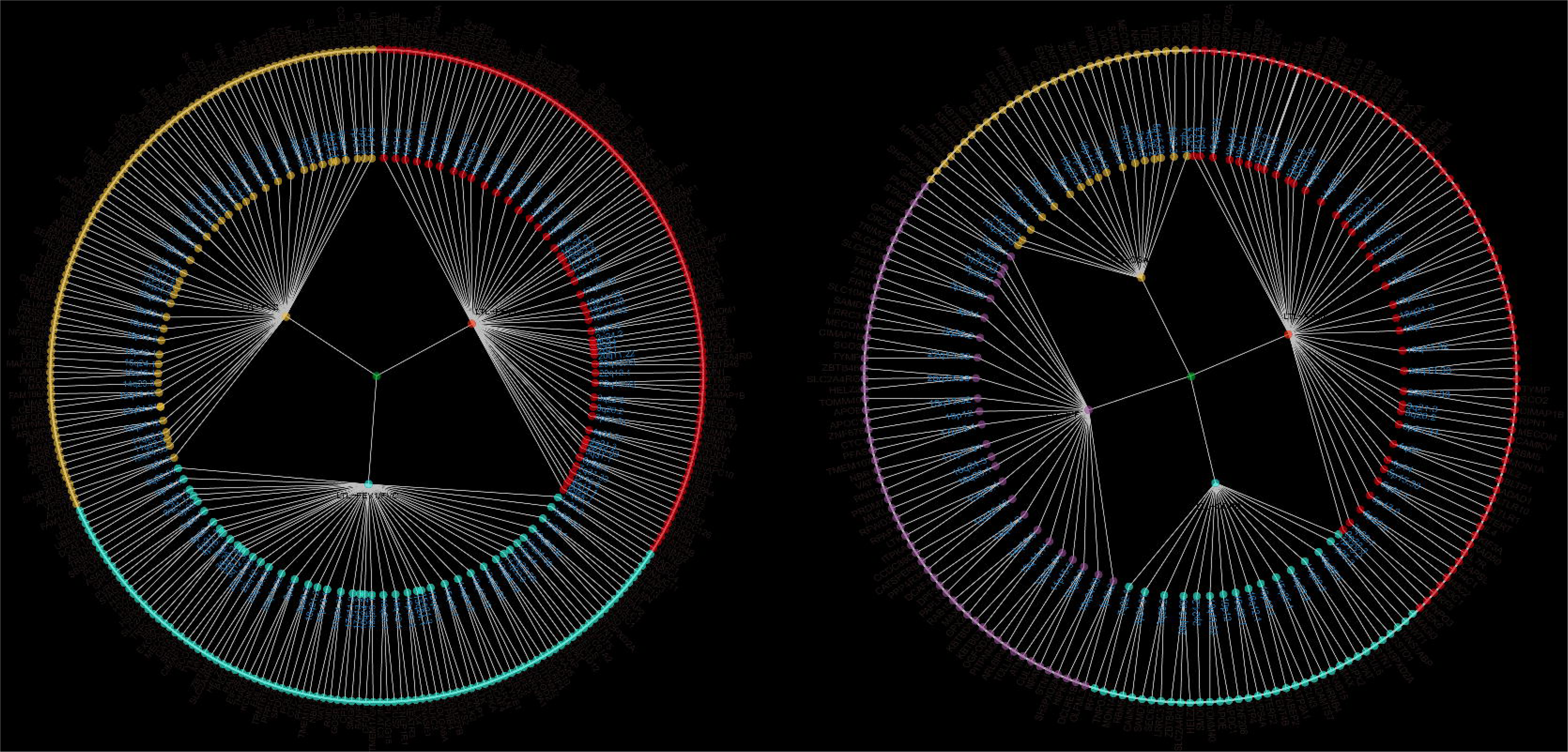
Overall landscape of pleiotropic associations between LTL and seven major respiratory health indicators. The circular dendrogram is centered on LTL (inner circle) and forms seven trait pairs with four chronic respiratory diseases (second circle, Asthma, COPD, IPF and OSA) and three lung function parameters (second circle, FEV1, FEV1/FVC and FVC). A total of 329 pleiotropic loci were identified in these trait pairs (third circle). A total of 382 significant pleiotropic genes (122 unique genes) were identified by MAGMA analysis. For trait pairs with more than three pleiotropic genes, we only show the top three pleiotropic genes in order of priority (fourth circle).

To identify the tissues in which the above SNPs exert their effects, we performed LDSC-SEG analysis to determine tissue and functional category enrichments for each trait. We found that LTL-associated SNPs were significantly enriched in EBV-transformed lymphocytes. For IPF, the top enriched tissues included EBV-transformed lymphocytes and skin (sun-exposed lower leg). Asthma-related SNPs showed significant enrichment in whole blood. Notably, previous studies have also reported Asthma-related enrichment in lung and skin tissues. Based on these findings, we selected five relevant tissues, EBV-transformed lymphocytes, lung, skin (both sun-exposed and not sun-exposed), and whole blood, for further tissue-specific gene mapping analyses (Supplementary Table 13).

We then conducted tissue-specific e-MAGMA analysis across the five selected tissues using GTEx v8 expression models. After Bonferroni correction, a total of 2,082 (462 Unique genes) significant tissue-specific genes were identified, of which 263 genes (56.93%) were associated with more than one trait pair (Supplementary Table 14). Among them, *STMN3* and *UCKL1*, both located in the 20q13.33 region, were significantly associated with all trait pairs. *STMN3* showed strong tissue specificity in skin (sun-exposed lower leg) and whole blood. *UCKL1*, an enzyme in the pyrimidine salvage pathway, was primarily enriched in whole blood. While *UCKL1* has been linked to various cancers[35][36], such as lung cancer, its involvement in CRDs, LF parameters, or LTL has not been previously reported. We also identified 19 tissue-specific genes associated with six trait pairs. Notably, *MPHOSPH9* and *MTRFR*, both located at 12q24.31, were consistently associated across these trait pairs, except for LTL-COPD. *MPHOSPH9* showed enrichment in both lung and skin tissues, suggesting a potential lung-specific role in LTL and CRD-related pathways. *MTRFR* was enriched predominantly in both sun-exposed and non-sun-exposed skin, implicating a skin-related mechanism in disease progression. To further prioritize likely causal genes, we performed transcriptome-wide association studies (TWAS), which identified 1,133 novel genes associated with LTL and 1,479 novel genes associated with respiratory health indicators (Supplementary Table 15). Furthermore, 1,808 (86.84%) tissue-specific genes identified by e-MAGMA were validated using FUMA eQTL mapping, providing strong support for their regulatory relevance (Supplementary Table 8).

In total, MAGMA and e-MAGMA jointly identified 222 pleiotropic genes (82 unique genes) (Figure 4). Among these, 26 genes were involved in more than half of the trait pairs, with enrichment primarily in the 12q24.31, 20q13.33, and 14q23.3 regions. Notable pleiotropic genes included *STN1*, *MPHOSPH9, MTRFR*, *MAX*, and *UCKL1*. While each gene was absent in one or two specific trait pairs (e.g., *STN1* in LTL-OSA), their repeated involvement across most trait pairs underscores their potential central role. For example, *MAX*, known for its role in cell proliferation and inactivation in small-cell lung cancer (approximately 6% of cases)[37], may also contribute to LTL regulation. Similarly, *MTHFR*, a key enzyme in the methionine-folate cycle, has been linked to Asthma susceptibility (particularly the C677T variant), although its mechanistic role in respiratory phenotypes remains to be clarified[38].

### Shared genetic mechanisms between LTL and respiratory health indicators

To better understand the biological functions and pathways underlying the genetic associations between LTL and respiratory health indicators, we performed gene set enrichment analysis on the MAGMA-mapped genes. This analysis identified 44 significant biological pathways (P < 7.45 × 10^−7^) (Supplementary Figure 4 and Supplementary Table 16). Among the top-ranked pathways, positive regulation of macromolecule biosynthetic processes and positive regulation of RNA metabolic processes were notably enriched in four trait pairs: LTL-Asthma, LTL-FEV1, LTL-FEV1/FVC, and LTL-FVC. Further examination of gene sets associated with LTL-Asthma and LTL-COPD revealed convergence on pathways related to telomere maintenance, including telomere lengthening and negative regulation of telomere maintenance. This observation was further supported by functional enrichment analysis of 82 overlapping pleiotropic genes identified via MAGMA and e-MAGMA using the Metascape database (Supplementary Table 17). Notably, genes such as *STN1*, *MAX*, and *MPHOSPH9* were extensively involved in these telomere-related pathways. Moreover, given that numerous clinical studies have linked premature aging to the development of COPD and Asthma, the abnormal shortening of LTL, may be a key mechanism triggering premature aging, which warrants further investigation.

### Causal inference between LTL and respiratory health indicators

Studies on horizontal pleiotropy have highlighted potential shared biological mechanisms between trait pairs, but genetic causal effects (i.e., vertical pleiotropy) remain largely unclear. To clarify the causal relationships between LTL and respiratory health indicators, we applied the latent causal variable (LCV) approach. Genetic causality was absent for most trait pairs based on the criteria of p > 7.14 × 10^−3^ and |GCP| < 0.6 (Supplementary Table 18A). Notably, only the LTL-IPF pair showed a p-value below the strict Bonferroni threshold, with the |GCP| value approaching 0.6 closely. This finding demonstrates that shorter LTL is causally associated with an increased risk of IPF. We further applied MRlap analysis, correcting for sample overlap, which supported a causal effect of short LTL on increased IPF risk (Supplementary Table 18B). Additionally, reverse MR analysis revealed a significant negative causal effect of genetically predicted COPD on LTL (*P* = 3.69 ×10^−5^). In summary, these findings provide partial evidence supporting vertical pleiotropy and highlight the need for a more nuanced understanding of the genetic architecture underlying LTL and respiratory health indicators.

## Discussion

This genome-wide pleiotropic association study investigates the shared genetic architecture between LTL and respiratory health indicators. Using a multi-level genomic analytical framework, we examined genetic correlations, genetic overlap, pleiotropic loci and genes, and bidirectional causal relationships. Our results revealed extensive genetic sharing that is not fully captured by genome-wide genetic correlation, underscoring a substantial basis of pleiotropy between LTL and respiratory health indicators. Specifically, we identified 27,885 pleiotropic variants and 82 unique pleiotropic genes, highlighting their potential roles in influencing susceptibility to LTL, CRDs and LF parameters. To further interpret these associations biologically, pathway enrichment analysis identified macromolecular biosynthesis, RNA metabolism, and telomere maintenance as key shared biological pathways underlying the genetic links between LTL and respiratory health indicators. Regarding potential causal effects (vertical pleiotropy), LCV analysis provided evidence of a negative causal association between LTL and IPF. Overall, our findings provide novel insights into the complex genetic interconnections between telomere biology and pulmonary health.

First, we observed significant genome-wide *r_g_s* for five out of seven trait pairs. Notably, LTL and IPF exhibited the strongest absolute genetic correlation, consistent with previous reports identifying IPF as the phenotype most strongly associated with short telomere defects. Consistently, LAVA and MiXeR analyses revealed widespread genetic overlap between LTL and respiratory health indicators, although the extent and direction of shared variants varied across trait pairs. For example, MiXeR analysis showed that more than half of LTL-associated variants also influenced OSA, despite the absence of a significant genome-wide genetic correlation between these traits. Supporting this, LAVA analysis identified 11 independent genomic regions (out of 1,822 tested) showing significant genetic correlations between LTL and OSA, with nearly 45% of these correlations being positively directed. Although previous studies on LTL changes in OSA patients have yielded conflicting results, the majority suggest that oxidative stress, hypoxia, and inflammation occurring in OSA patients may contribute to LTL shortening. In contrast, the genetic signals shared between LTL and COPD displayed a high degree of overlap with consistent effect directions. In summary, these findings underscore the importance of capturing the full spectrum of polygenic overlap, regardless of effect direction, to better understand the shared genetic basis between LTL and respiratory health indicators.

Given that both vertical (causal) and horizontal (biological) pleiotropy underlie the shared genetic basis of complex polygenic traits, we applied multiple analytical strategies to dissect their respective contributions. Here, we focus on horizontal pleiotropy, identifying 27,885 pleiotropic variants associated with LTL and respiratory health indicators via PLACO. Many of these variants appear to influence aging-related biological pathways in opposite directions, suggesting that shared molecular mechanisms may contribute to divergent phenotypic outcomes. Functional annotation revealed that pleiotropic variants shared across six or more trait pairs predominantly localized to chromosomal regions such as 12q24.31, 20q13.33, 3q26.2, 14q23.3, and 10q24.33. For example, *MPHOSPH9* (12q24.31), also known as matrix metalloproteinase 9 (*MMP9*), encodes an endopeptidase implicated in extracellular matrix remodeling and lung pathology, including fibrosis[39]. Elevated *MMP9* expression promotes pulmonary fibrosis by degrading collagen and elastin in the lung interstitium, disrupting airway homeostasis[40][41]. While *MMP9*’s role in lung diseases is well established, its genetic association with LTL remains unexplored and warrants further study. Another pleiotropic gene, *MAX* (14q23.3), a tumor suppressor forming heterodimers with Myc, regulates transcription of hTERT, essential for telomerase activation and cell immortalization[42][43]. Mutations in *MAX* have been linked to several lung cancers[37], though its role in non-malignant lung diseases and pulmonary function remains unclear, highlighting the need for further functional studies.

To enhance gene mapping reliability, we employed two complementary strategies and identified five pleiotropic genes shared across up to six trait pairs, including *STN1*, *MPHOSPH9*, *MTRFR*, *MAX*, and *UCKL1*. *STN1* (10q24.33), encoding a protein critical for telomere replication and capping, was associated with six respiratory phenotypes, excluding LTL-OSA. *STN1* recruits DNA polymerase α-primase to telomeres, maintaining chromosome integrity[32][33]. Previous studies linked *STN1* to LTL, IPF, COPD, and lung cancer[34][44][45]. Our analysis further uncovers its pleiotropic role across Asthma, and LF. *UCKL1*, involved in pyrimidine salvage and regulation of the antioxidant Nrf2 pathway, may contribute to lung cellular defense mechanisms[46]. Although direct evidence linking *UCKL1* to CRDs is lacking, dysregulated Nrf2 signaling in COPD alveolar macrophages suggests a potential pathogenic mechanism[47]. *MTRFR* (*C12orf65*), a mitochondrial peptide release factor involved in organellar RNA repair, has shown genetic associations with multiple respiratory health indicators, warranting further investigation into its potential role in respiratory function[48][49].

Pathway enrichment analyses of pleiotropic loci shared between LTL-LF, and LTL-Asthma revealed overlapping pathways involved in macromolecular biosynthesis and RNA metabolism. These results align with epidemiological evidence linking frequent Asthma exacerbations to progressive LF decline[50]. Functional annotation also highlighted enrichment in pathways negatively regulating telomere elongation and maintenance, consistent with telomere shortening-induced cellular senescence and apoptosis. Accelerated telomere attrition correlates with impaired alveolar gas exchange in COPD[51]. Knockout of telomerase reverse transcriptase and telomerase RNA component leads to alveolar cell senescence and lung inflammation[52]. Airway smooth muscle cells from Asthma patients similarly display elevated senescence markers[53], suggesting that shared genetic factors modulate telomere biology and contribute to respiratory disease pathogenesis. Collectively, these findings elucidate the pleiotropic genetic architecture linking telomere biology and respiratory health indicators, providing a foundation for future functional studies and potential therapeutic targeting.

At the level of vertical pleiotropy, we employed the LCV approach and MRlap to explore potential causal associations between LTL and respiratory health indicators. Consistent with the results reported by Duckworth *et al.*[15], our study supports a causal relationship whereby longer LTL reduces the risk of IPF. However, discrepancies remain concerning the causal relationship between COPD risk and LTL[15][54]. These inconsistencies may arise from inherent limitations of two-sample MR, including potential sample overlap, which warrants cautious interpretation of prior MR findings. Overall, our LCV and MRlap analyses not only extend previous MR studies but also suggest that the shared genetic architecture between LTL and respiratory health indicators is predominantly driven by horizontal pleiotropy rather than vertical pleiotropy.

This study has several limitations. First, the genetic analysis methods rely on stringent assumptions, such as the absence of the independence of genetic instruments from confounders, which may introduce bias despite our efforts to enhance robustness through complementary analytical strategies. Potential residual confounding cannot be entirely excluded. Second, our analyses were restricted to GWAS data from individuals of European ancestry, primarily due to differences in sample size availability and LD patterns across populations, which may limit the generalizability of our findings. Future cross-ancestry studies are essential to evaluate the consistency of these genetic associations across diverse populations. Third, although we identified shared biological pathways potentially linking LTL with respiratory health indicators, the exact pathophysiological mechanisms remain unclear. Further experimental and functional studies are needed to elucidate the molecular mechanisms underlying these genetic associations and to confirm their biological relevance.

## Conclusion

This study investigates the polygenic overlap between LTL and respiratory health indicators, revealing a stronger genetic correlation than previously reported. Our findings highlight shared molecular pathways, particularly those involving RNA metabolism and telomere maintenance, that underlie the common genetic architecture of respiratory health indicators. In conclusion, this work not only reinforces the genetic link between telomere-mediated cellular senescence and chronic lung diseases but also provides novel insights into potential biomarkers and therapeutic targets for the assessment and management of chronic lung disease patients.

## Acknowledgements

This work was funded by the Natural Science Foundation of China Excellent Young Scientists Fund (Overseas) (Grant No. K241141101), Guangdong Basic and Applied Basic Research Foundation for Distinguished Young Scholars (Grant No. 2024B1515020047), Shenzhen Basic Research General Projects of Shenzhen Science and Technology Innovation Commission (Grant No. JCYJ20230807093514029), National Natural Science Foundation of China (Grant No. 82470452), Natural Science Foundation of Xinjiang Uygur Autonomous Region (Grant No. 2024D01D15) (To Y.F.), Shenzhen Science and Technology Program, Shenzhen, China (Grant No. GJHZ20240218111401002), National Science and Technology Major Project (Grant No. 2023ZD0505902) (To Jin-Song Bian), and Center for Computational Science and Engineering at Southern University of Science and Technology. The funder had no role in the design, implementation, analysis, interpretation of the data, approval of the manuscript, and decision to submit the manuscript for publication.

## Footnotes

Author contributions: P.Z. and Y.F. conceptualized and supervised this project and wrote the manuscript. P.Z., L.Z., J.Q., and Y.F.. performed the main analyses and wrote the manuscript. P.Z., Y.S., Y.N., and Y.Y. performed the statistical analysis and assisted with interpreting the results. M.Y., X.H. and J.G. provided expertise in telomere biology and respiratory health. All authors provided intellectual content and approved the final version of the manuscript.

## Data availability

GWAS summary statistics on LTL are available at https://figshare.com/s/caa99dc0f76d62990195. GWAS summary statistics on IPF, OSA, FEV1, FEV1/FVC, and FVC are available at the GWAS Catalog (GCST90133317, GCST009541, GCST007432, GCST007431, and GCST007429). GWAS summary statistics on Asthma and COPD are publicly available for download at the Global Biobank Meta-analysis Initiative website: https://www.globalbiobankmeta.org/resources.

## Code availability

All software (and version, where applicable) used to conduct the analyses in this paper are freely available online: LDSC (v1.0.1; https://github.com/bulik/ldsc), MiXeR (v1.3; https://github.com/precimed/mixer), LAVA (v0.1.0; https://github.com/josefin-werme/LAVA), PLACO (v0.1.1; https://github.com/RayDebashree/PLACO), FUMA (v1.5.4; http://fuma.ctglab.nl/), COLOC (v5.2.1; https://github.com/chr1swallace/coloc), MAGMA (v.1.08; https://ctg.cncr.nl/software/magma), e-MAGMA (https://github.com/eskederks/eMAGMA-tutorial), TWAS (http://gusevlab.org/projects/fusion/), LCV (https://github.com/lukejoconnor/LCV), MRlap (https://github.com/n-mounier/MRlap/), and R (v.4.1.3; https://www.r-project.org/).

---

**Supplementary Figure 4:**
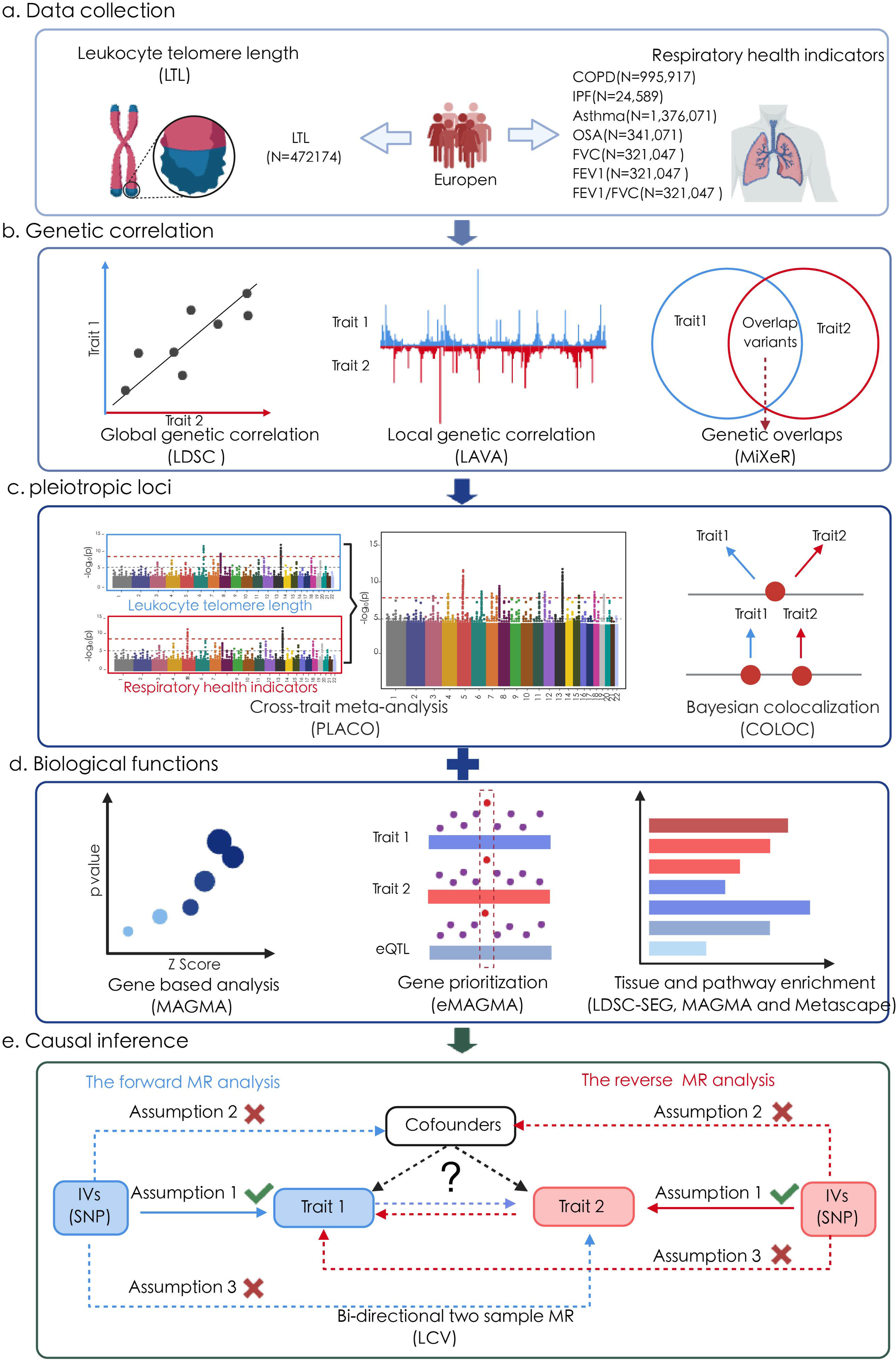
MAGMA pathway enrichment of LTL and seven major respiratory health indicators. The x-axis reflects the phenotype pairs, and the y-axis reflects the pathways from the Molecular Signatures Database dataset. The color gradient represents the magnitude of the value, and different colors correspond to different value ranges. Dark red represents higher values.

